# Immune response to the mRNA COVID-19 vaccine in hemodialysis patients: cohort study

**DOI:** 10.1101/2023.01.19.23284792

**Authors:** Yi-Shin Chang, Kai Huang, Jessica M Lee, Christen L Vagts, Christian Ascoli, Md-Ruhul Amin, Mahmood Ghassemi, Claudia M Lora, Russell Edafetanure-Ibeh, Yue Huang, Ruth A Cherian, Nandini Sarup, Samantha R Warpecha, Sunghyun Hwang, Rhea Goel, Benjamin A Turturice, Cody Schott, Montserrat Hernandez, Yang Chen, Julianne Joregensen, Wangfei Wang, Mladen Rasic, Richard M Novak, Patricia W Finn, David L Perkins

## Abstract

**Background:** End-stage renal disease (ESRD) patients experience immune compromise characterized by complex alterations of both innate and adaptive immunity, and results in higher susceptibility to infection and lower response to vaccination. This immune compromise, coupled with greater risk of exposure to infectious disease at hemodialysis (HD) centers, underscores the need for examination of the immune response to the COVID-19 mRNA-based vaccines.

**Methods:** A transcriptomic analysis of the immune response to the Covid-19 BNT162b2 mRNA vaccine was assessed in 20 HD patients and cohort-matched controls. RNA sequencing of peripheral blood mononuclear cells (PBMCs) was performed longitudinally before and after each vaccination dose for a total of six time points per subject. Anti-spike antibody levels were quantified prior to the first vaccination dose (V1D0) and seven days after the second dose (V2D7) using anti-Spike IgG titers and antibody neutralization assays. Anti-spike IgG titers were additionally quantified six months after initial vaccination. Clinical history and lab values in HD patients were obtained to identify predictors of vaccination response.

**Results:** Transcriptomic analyses demonstrated differing time courses of immune responses, with predominant T cell activity in controls one week after the first vaccination dose, compared to predominant myeloid cell activity in HD at this time point. HD demonstrated decreased metabolic activity and decreased antigen presentation compared to controls after the second vaccination dose. Anti-spike IgG titers and neutralizing function were substantially elevated in both controls and HD at V2D7, with a small but significant reduction in titers in HD groups (p < 0.05). Anti-spike IgG remained elevated above baseline at six months in both subject groups. Anti-spike IgG titers at V2D7 were highly predictive of 6-month titer levels. Transcriptomic biomarkers after the second vaccination dose and clinical biomarkers including ferritin levels were found to be predictive of antibody development.

**Conclusion:** Overall, we demonstrate differing time courses of immune responses to the BTN162b2 mRNA COVID-19 vaccination in maintenance hemodialysis subjects (HD) comparable to healthy controls (HC) and identify transcriptomic and clinical predictors of anti-Spike IgG titers in HD. Analyzing vaccination as an in vivo perturbation, our results warrant further characterization of the immune dysregulation of end stage renal disease (ESRD).

**Funding:** F30HD102093, F30HL151182, T32HL144909, R01HL138628

This research has been funded by the University of Illinois at Chicago Center for Clinical and Translational Science (CCTS) award UL1TR002003

## Background

End-stage renal disease (ESRD) is the most advanced stage of chronic kidney disease (CKD), with prevalence in the U.S. reaching 809,000 in 2019 ^1^. The most used form of renal replacement therapy for ESRD patients in the U.S. is hemodialysis (HD). Despite significant improvements in hemodialysis technology, the mortality rate in ESRD patients is still as high as 20% annually ^2^, with infections being the most common cause of hospitalization and mortality after cardiovascular disease ^3^. The immunocompromised state of ESRD is characterized by simultaneous immunodepression due to the impact of uremic milieu on immunocompetent cells and immunoactivation due to the accumulation of proinflammatory cytokines ^3^. There are alterations to both innate and adaptive immunity, including elevated levels of mannose-binding lectin ^4^, impaired maturation of monocytes and dendritic cells ^4,5^, increased B cell apoptosis ^6^, and decreased T-cell proliferation with elevated Th1/Th2 ratio^7^.

Studies of genome-wide expression (i.e. transcriptome) profiles of peripheral blood mononuclear cells in ESRD demonstrate a complex picture of immune alterations. One study found upregulation of genes involved in the complement and oxidative metabolism pathways, and downregulation of genes associated with the clathrin-coated vesicle endosomal pathway and T-cell receptor signaling ^10^. Two other studies have demonstrated impaired expression of genes involved in oxidative phosphorylation and mitochondrial function ^11,12^. A study identifying a group of inflammatory genes playing a causative role in oxidative stress in dialysis patients showed unique gene expression alterations in maintenance HD patients compared to un-dialyzed CKD patients and compared to patients undergoing peritoneal dialysis ^13^. These studies indicate a range of immune pathways that may impair vaccination response, and further suggest that dialysis leads to unique immune profile alterations. These alterations may lead to higher susceptibility to infection and lower response to vaccination ^8^. For example, while more than 90% of patients without CKD develop protective antibodies against HBV after vaccination, only 50-60% of patients with ESRD seroconvert. There have also been higher vaccination failure rates demonstrated against influenza virus, Clostridium tetani, and Corynebacterium diphtheriae in ESRD ^9^.

Understanding the immune response to vaccines in ESRD is particularly important for the new mRNA vaccines developed in response to the COVID-19 pandemic. The COVID-19 mRNA-based vaccines, BNT162b2 and mRNA-1273, have proven to be efficacious in non-immunocompromised individuals, with initial reports showing 95% and 94.1% reduction of COVID-19 disease in recipients ^14,15^. However, certain immunosuppressed populations remain at risk of infection. Given the widespread transmission of COVID-19, detailed assessments of degree, duration, and determinants of immune protection conferred by these vaccines are vitally needed in immunocompromised patient populations including those with ESRD.

While recent studies of the SARS-CoV-2 BTN162b2 vaccine in HD demonstrate high levels of seroconversion ranging from 84-96% ^16–19^, they also demonstrate quantitatively reduced SARS-CoV-2 IgG antibodies. We posit that characterization of the transcriptomic underpinnings of antibody titer development on a continuous scale may identify biomarkers for weaker or less durable immune protection in this population. Furthermore, transcriptomic analyses may identify strategies for the development of new, effective vaccines against other infectious diseases for this population. Thus, we characterized the immune response of the HD population to the COVID-19 mRNA-based BNT162b2 vaccine using mRNA sequencing, anti-spike antibody ELISA and neutralization titers across multiple time points. We then integrated these data to identify transcriptomic and clinical determinants of the humoral immune response in HD patients.

## Methods

### Study population and sample acquisition

The study was approved by the University of Illinois at Chicago IRB (#2018-1038) Ethics Review Committee. Maintenance HD patients undergoing vaccination with the BNT162b2 mRNA COVID-19 vaccine in February 2021 were recruited from the outpatient HD unit at the University of Illinois Hospital (UIH) in Chicago, IL. Control subjects consisted of UIH employees undergoing BNT162b2 mRNA COVID-19 vaccination at UIH from December 2020 to January 2021 with no self-reported history of kidney disease or immune disorders. A subset of control subjects matched for age, gender, and COVID-19 history was also analyzed for this study. Blood was collected at 0 – 48 hours prior to and at multiple time points after both the first (V1) and second vaccination doses (V2), which were administered three weeks apart. Control samples were collected prior to each vaccination dose (D0) and at one day (D1) and seven days (D7) after each dose, corresponding to six time points: V1D0, V1D1, V1D7, V2D0, V2D1, V2D7. Blood was collected from HD subjects prior to each vaccination dose and at two days (D2) and seven days after each dose, corresponding to six time points: V1D0, V1D2, V1D7, V2D0, V2D2, V2D7. A final blood sample was drawn six months after initial vaccination (M6) for measurement of antibody titers, prior to additional vaccination doses. Serum and peripheral blood mononuclear cells (PBMCs) were extracted within two hours of blood collection, then stored at −80°C. PBMCs were extracted using density gradient centrifugation at 400g with Ficoll-Paque PLUS. The extracted buffy coat was stored in RNAlater (Invitrogen).

### Clinical and Demographic Characterization

Demographic and clinical data was collected from the electronic health record (EHR) for HD subjects, including medical diagnoses, medications, and laboratory values. Laboratory values included monthly SARS-CoV-2 test results, as well as urea reduction ratio (URR, a measure of dialysis adequacy), hemoglobin (Hgb), ferritin, transferrin saturation, albumin levels, white blood cell (WBC) count and WBC differential counts obtained during standard of care monthly blood draws for the three months preceding vaccination. Within our analyses, ferritin was coded as either low risk (200ng/ml – 1200 ng/ml) or high risk (<200 ng/ml or >1200 ng/ml), since ferritin levels 200ng/ml – 1200 ng/ml have been shown to be associated with lowest all-cause mortality in HD patients ^20^. Baseline clinical lab values were calculated as the median of three lab values across the three months prior to vaccination. Demographic and clinical data was collected from a medical questionnaire at time of consent for control subjects, and included medical history, medications, and self-reported prior SARS-CoV-2 positive test results.

### RNA extraction and RNA Sequencing (RNAseq)

RNA sequencing was performed on PBMCs at all V1 and V2 time points for all subjects for whom RNA libraries were successfully built at ≥ 5 time points. PBMCs stored in RNAlater were thawed and diluted 1:1 with 1X phosphate buffered saline. The mixture was then pelleted and RNA was extracted using the PureLink RNA Mini kit (Invitrogen). DNase treatment to remove genomic DNA contamination was performed using either the PureLink DNase kit or DNA-free DNA Removal Kit (Invitrogen). Purified RNA in sterile water was stored at −80°C. Each RNA sample was quantified using the Qubit RNA High Sensitivity kit (Invitrogen) and Bioanalyzer RNA Pico kit (Agilent) with RIN>=8.

For library construction, 50ng of RNA from each sample was aliquoted in 96 well plates. Libraries were generated using the NEBNext Ultra II Directional RNA Library Prep Kit for Illumina with the optional NEBNext Poly(A) mRNA Magnetic Isolation Module (New England BioLabs). Each individual sample library was barcoded during PCR amplification using unique dual indexed i5 and i7 primers from the NEBNext Multiplex Oligos for Illumina kit. Each sample library was quantified using the Qubit DNA High Sensitivity kit and Bioanalyzer DNA High Sensitivity kit. Samples were then pooled and sequenced using the MiSeq Nano V2 kit (Illumina) to check read proportions between samples. Samples with lower-than-expected percentage of reads detected were supplemented with an additional spike-in of sample library to the main pool. The supplemented pooled library was sequenced again using the MiSeq Nano V2 kit to verify adequate adjustment. The finalized library was sequenced using a NovaSeq S2 flow cell configured for 75bp paired end output.

### Differential Gene Expression Analysis

Raw demultiplexed reads were filtered using fastp to remove adapters and short reads ^21^. Trimmed reads were then quantified using the Salmon pipeline with an hg38 reference transcriptome index ^22^. Quantified data was imported into R using the tximeta package ^23^ to convert Salmon quantification and index data to a count matrix. Transcript names were extracted and matched using Entrez IDs with the AnnotationHub package ^23^. This finalized count matrix was then imported into a DESeqDataset object and normalized using the variance stabilizing transformation in DESeq2.

The *DESeq2* R package was used to identify genes that were differentially expressed at each time point after vaccination for each subject group. Specifically, we implemented a design incorporating group-specific condition effects with individual subjects nested within groups. We performed the classical *Deseq2* workflow of estimation of size factors, estimation of dispersion, and negative binomial GLM fitting for β_i_ and Wald statistics, increasing the maximum number of iterations for estimation of the negative binomial distribution to 500. We then generated contrasts to obtain differentially expressed genes for controls at V1D1 and V1D7 (compared to V1D0), and at V2D1 and V2D7 (compared to V2D0). Differentially expressed genes for HD were similarly obtained at V1D2 and V1D7 (compared to V1D0), and at V2D2 and V2D7 (compared to V2D0). We also directly compared gene expression between controls and HD at V1D7 and at V2D7. The significance threshold to determine differential expression was FDR-adjusted (p < 0.05).

### Anti-Spike (trimer) IgG Titer Quantification

The Human SARS-CoV-2 Spike (Trimer) IgG ELISA Kit from Invitrogen was used to quantitate IgG to the SARS-CoV-2 spike protein in serum samples at V1D0, V2D7, and M6 time points. All samples were initially diluted 1:100 (in addition to the 1:10 assay buffer dilution on the 96-well plate) and assayed in duplicate, with two-fold serial dilution of the 150,000 units/mL standard control in duplicate for relative quantification. Absorbance at 450 nm was quantified using a *Spark*® multimode microplate reader. Samples that produced signals greater than the upper limit of the standard curve were diluted 1:2000 and assayed again. IgG concentration was calculated by fitting four-parameter logistic curves to the standard controls and taking the average concentrations of duplicates.

### Antibody Neutralization Assays

Neutralization assays were performed on serum samples from V1D0 and V2D7 using SARS-CoV-2 pseudotyped virus (pseudovirus). To produce pseudoviruses, an expression plasmid bearing codon-optimized SARS-CoV-2 full length S plasmid was co-transfected into HEK293T cells using the SARS-CoV-2 Spike-pseudotyped lentiviral particle Kit (BEI # R-52948). The cell supernatants were collected 72h after transfection, divided into aliquots and cryopreserved at −80 °C.

To titrate the pseudovirus, 5×10^3^ 293T-ACE2 cells were seeded per well in a 96-well plate in DMEM containing 10% FBS and 1% penicillin streptomycin. Twenty-four hours later, the pseudovirus was diluted 1:10, followed by five-fold serial dilutions for a total of nine dilutions, with each dilution performed in six replicate wells. After incubation at 37 °C and 5% (vol/vol) CO2 for 72h, the luciferase substrate was added to the 96-well plate for chemiluminescence detection. The 50% tissue culture infectious dose (TCID50) of the pseudovirus was calculated according to the Reed-Muench method in the titration macro template ^24^.

Neutralization activity against SARS-2-CoV was measured in a single-round-of-infection assay with pseudoviruses as previously described ^25^. 5×10^3^ 293T-ACE2 cells were seeded per well in a 96-well plate. Twenty-four hours later, serial dilutions of the serum samples were performed, incubated for one hour at 37 °C with ∼1000 TCID50/ml of pseudovirus, then added to monolayers of ACE2-overexpressing 293T cells in quadruplicate. The cell control with cells alone and the virus control (VC) with pseudovirus were set up in each plate. The target cells were incubated for 65h-72h at 37 °C and 5% (vol/vol) CO2. Fifty μL of Bright-Glo, reconstituted following manufacturer’s instructions, was added to each well of the 96-well plate and incubated for five minutes at room temperature. The 96-well plate was read by a 96-well luminescence plate reader (Tecan Genius Pro plate reader) ^26^. Percent neutralization was calculated as 100*([Virus-only control] – [Virus plus serum])/[Virus-only control], and neutralizing titer levels are reported as the serum dilution required to achieve 50% neutralization (50% inhibitory dilution [ID50]) ^27^. The input dilution of serum was 1:20, thus 20 is the lower limit of quantification.

### BTM module enrichment analysis

Gene set enrichment analysis was performed for each contrast generated in the DESeq2 analysis above using blood transcription module (BTMs) gene sets ^28^. BTMs with FDR-adjusted p < 0.05 were considered significantly enriched. Enriched BTMs were further characterized using the distribution of Wald statistics of membership genes from DESeq2. To summarize BTM analyses, BTMs were categorized into different families: B cells, cell cycle, dendritic cell/antigen presentation, type I interferon (IFN type I), myeloid activity/inflammation/ T/NK cells, and “others” ^29^. The percentage of BTMs in each BTM family with significant enrichment at each time point was then quantified over time.

### Statistical analysis of antibody response

To determine the effect of vaccination on anti-spike IgG titers at V2D7 and M6, Kruskal-Wallis tests were performed separately for HD subjects and controls. For each group, anti-spike IgG titer levels were compared to assess for the significant effect of time (V1D0, V2D7, M6), and Wilcoxon rank sum tests were performed with FDR correction to assess significant differences between each pair of time points (V2D7 vs. V1D0, M6 vs. V1D0, M6 vs. V2D7). To determine the effect of vaccination on antibody neutralization activity (ID50) at V2D7, Wilcoxon rank sum tests were performed for each group to compare V2D7 vs. V1D0.

Linear models were constructed to establish the effect of prior SARS-CoV-2 infection and subject group on anti-spike IgG titer development at V2D7 and M6 and neutralization activity at V2D7. Specifically, log-transformed V2D7 anti-spike IgG titers or V2D7 neutralization activity (ID50) were modeled as the dependent variable, with subject group (HD or controls), log-transformed V1D0 anti-spike IgG titers or V1D0 neutralization activity (ID50), gender, age, race, and ethnicity as independent predictors. To determine predictors of anti-spike IgG at six months, a linear model was constructed with the log-transformed M6 anti-spike IgG titers as the dependent variable, and V2D7 anti-spike IgG titers, SARS-CoV-2 history, gender, age, race, and ethnicity as independent predictors.

### Identification of BTM and clinical predictors of Ab response in HD

BTM predictors of antibody response in HD were identified by first calculating a representative expression level of each BTM per sample, which we will refer to as the eigengene. Specifically, the first principal component of each BTM was calculated using DESeq2-derived variance-stabilized gene counts from each module’s member genes across the HD V1 time points, and then across the HD V2 time points. Signs (positive or negative) were assigned to the eigengenes such that samples with higher expression of member genes in a BTM would be given a positive sign, while those with lower overall gene expression would be given a negative sign. This was accomplished for each BTM by (1) computing the median gene expression level across membership genes in a given BTM for each sample, (2) computing the Pearson correlation between the eigengene of the BTM and the median gene expression level across all samples, and (3) multiplying the eigengene of the BTM by −1 if the correlation was negative.

Subsequently, we constructed linear models with log-transformed anti-spike IgG at V2D7 as the dependent variable and change in BTM eigengene expression after vaccination as the independent variable, controlling for SARS-CoV-2 history. Separate models were constructed for each BTM that was enriched at each time point after vaccination in HD (V1D2 vs V1D0, V1D7 vs V1D0, V2D2 vs V2D0, V2D0 V2D7). Change in BTM expression was calculated as the BTM eigengene after vaccination minus the BTM eigengene before vaccination. P-values were FDR-adjusted across number of enriched BTMs per time point.

Additionally, baseline clinical laboratory values predictive of antibody response in the HD subjects were identified. Linear models were separately constructed using URR, ferritin (high risk vs low risk), transferrin saturation, hemoglobin, and WBC count to predict log-transformed anti-spike IgG at V2D7 and M6.

Finally, clinical laboratory values responding to vaccination that predicted antibody titer response in the HD subjects were identified. Linear models were separately constructed using log-fold change (LFC) from baseline measurements of ferritin (continuous instead of binarized low- and high-risk), transferrin saturation, and WBC count to predict log-transformed anti-spike IgG titers at V2D7 and M6.

## Results

### Demographic and Clinical Characterization

Demographic and clinical data of the 20 maintenance hemodialysis (HD) and controls (HC) are summarized in **Table 1**. The racial distribution differed between cohorts with more Black/African American subjects in the HD cohort. The cohorts were otherwise demographically similar. The subjects within the HD cohort had significantly more comorbidities, most notable of which include type 2 diabetes mellitus (T2DM), hypertension (HTN), dyslipidemia, and other cardiovascular conditions. The most common causes of renal failure were T2DM and HTN, with a minority of cases attributed to anatomic defects (reflux uropathy) and autoimmune conditions (systemic lupus erythematous and idiopathic thrombocytopenic purpura).

**Table 1.** Demographic and clinical data for maintenance hemodialysis and control subjects.

|  | Hemodialysis | Control | P-val |
| --- | --- | --- | --- |
| **Total # of subjects** | 20 | 20 |  |
| **Gender** |  |  |  |
| **Male** | 11 | 10 | 1.0 |
| **Female** | 9 | 10 | 1.0 |
| **Age (mean (sd))** | 54 (12) | 54 (13) | 0.98 |
| **Race/Ethnicity** |  |  |  |
| **Black/African American** | 10 | 3 | 0.041 |
| **Asian/Pacific Islander** | 1 | 2 | 1.0 |
| **White/Caucasian** | 2 | 8 | 0.067 |
| **Hispanic/Latinx** | 7 | 6 | 1.0 |
| **Other** | 0 | 1 | 1.0 |
| **BMI, kg/m<sup>2</sup> (mean (sd))** | 27.8 (5.1) | 28.7 (6.4) | 0.61 |
| **Medical History** |  |  |  |
| **Diabetes** | 11 | 1 | 0.0012 |
| **Hypertension** | 18 | 4 | < 0.001 |
| **Other CV disease\*** | 9 | 0 | 0.0012 |
| **Dyslipidemia** | 10 | 0 | < 0.001 |
| **Autoimmune disease\*\*** | 3 | 0 | 0.23 |
| **Immunosuppression\*\*\*** | 1 | 0 | 1.0 |
| **Active Malignancy\*\*\*\*** | 1 | 0 | 1.0 |
| **Positive COVID-19 History** | 8 | 5 | 0.5 |

\* Includes coronary artery disease (CAD), congestive heart failure (CHF), atrial fibrillation (AF), peripheral vascular disease (PVD), and cerebral vascular accident (CVA)
\*\* includes systemic lupus erythematosus (SLE), immune thrombocytopenic purpura (ITP), microscopic polyangiitis (MPA)
\*\*\* hydroxychloroquine
\*\*\*\* defined as malignancy requiring treatment in the last six months; one patient with papillary thyroid cancer requiring thyroidectomy, no systemic treatment required

There were eight HD subjects who previously tested positive for SARS-CoV-2, with positive test dates ranging from 7 months to four weeks preceding vaccination. Five control subjects self-reported a prior positive SARS-CoV-2 test, with positive test dates ranging from 8 months to four weeks preceding vaccination. Detailed clinical characterization of HD subjects is summarized in **Table 2**. Notable laboratory data includes an elevated ferritin from normal (with high population variance), and anemia.

**Table 2.** Baseline clinical lab values for maintenance hemodialysis patients.

|  | Normal range | Mean (SD) |
| --- | --- | --- |
| **Kidney/HD status** |  |  |
| **Urea Reduction Ratio (URR)** | - | 0.74 (0.052) |
| **Months on HD** | - | 46 (44) |
| **Iron** |  |  |
| **Ferritin (ng/ml)** | 10 - 259 | 838 (550) * |
| **% Transferrin saturation** | 25 - 50 | 38 (13) |
| **Albumin** | 3.4 – 5 | 4.1 (0.40) |
| **CBC** |  |  |
| **WBCs (k/ul)** | 3.9 - 12 | 6.0 (2.1) |
| **Hgb (g/dl)** | 13.2 – 18 | 10.5 (1.5) * |
| **Lymphocytes (k/ul)** | 1.3 - 4.2 | 1.5 (0.7) |
| **Neutrophils (k/ul)** | 1.3 - 7.5 | 3.7 (1.5) |
| **Monocytes (k/ul)** | 0.4 - 1 | 0.5 (0.2) |
| **Eosinophils (k/ul)** | 0.2 - 0.5 | 0.2 (0.2) |

\* indicates value outside of normal range

All subjects received two BTN162b2 vaccination doses with the second dose (V2) administered three weeks after the first (V1). Anti-spike IgG binding and neutralizing assay data were obtained for all subjects prior to V1 (V1D0) and seven days after V2 (V2D7). RNA sequencing data was obtained for all control subjects prior to each vaccination dose (D0), and at one day (D1) and seven days (D7) after each dose, corresponding to six time points: V1D0, V1D1, V1D7, V2D0, V2D1, V2D7. One control subject is missing V2D0 data, and one is missing V2D1 data. RNA sequencing data was obtained for 12 HD subjects prior to each vaccination dose, and at two days (D2) and seven days after each dose, corresponding to six time points: V1D0, V1D2, V1D7, V2D0, V2D2, V2D7. Two HD subjects are missing V2D2 data. Sequencing data was not obtained for subjects with fewer than five time points of successfully constructed RNA libraries, due to time points without sample collection or failure to extract high quality mRNA from PBMCs. Six-month follow-up (M6) anti-Spike IgG binding titers were obtained for 15 HC subjects and 19 HD subjects. One HD subject tested positive for SARS-CoV-2 14 days after the second vaccination dose, demonstrating mild symptoms. None of the other subjects reported SARS-CoV-2 infection up to 6 months follow up after the second vaccination.

### Differential Gene Expression Analysis

To characterize the molecular basis of immune responses to vaccination in HC and HD, we performed differential gene expression analyses of the PBMC RNA sequencing data. There are substantially more differentially expressed genes (DEGs) in response to V2 compared to V1, and at D1 and D2 post-vaccination compared to D7 (Figure 1). For HC, the largest number of DEGs is found at V2D1, indicating the most transcriptional activity immediately after the 2^nd^ vaccine dose, followed by V2D7, V1D1, and V1D7. HD follows a similar pattern, with the largest number of DEGs found at V2D2, followed by V2D7, V1D2, and V1D7. Notably, HD subjects with no SARS-CoV-2 history (n = 6) have substantially lower numbers of DEGs than HD subjects with positive SARS-CoV-2 history (n = 6) at each time point, and particularly at V2 time points.

**Figure 1.**
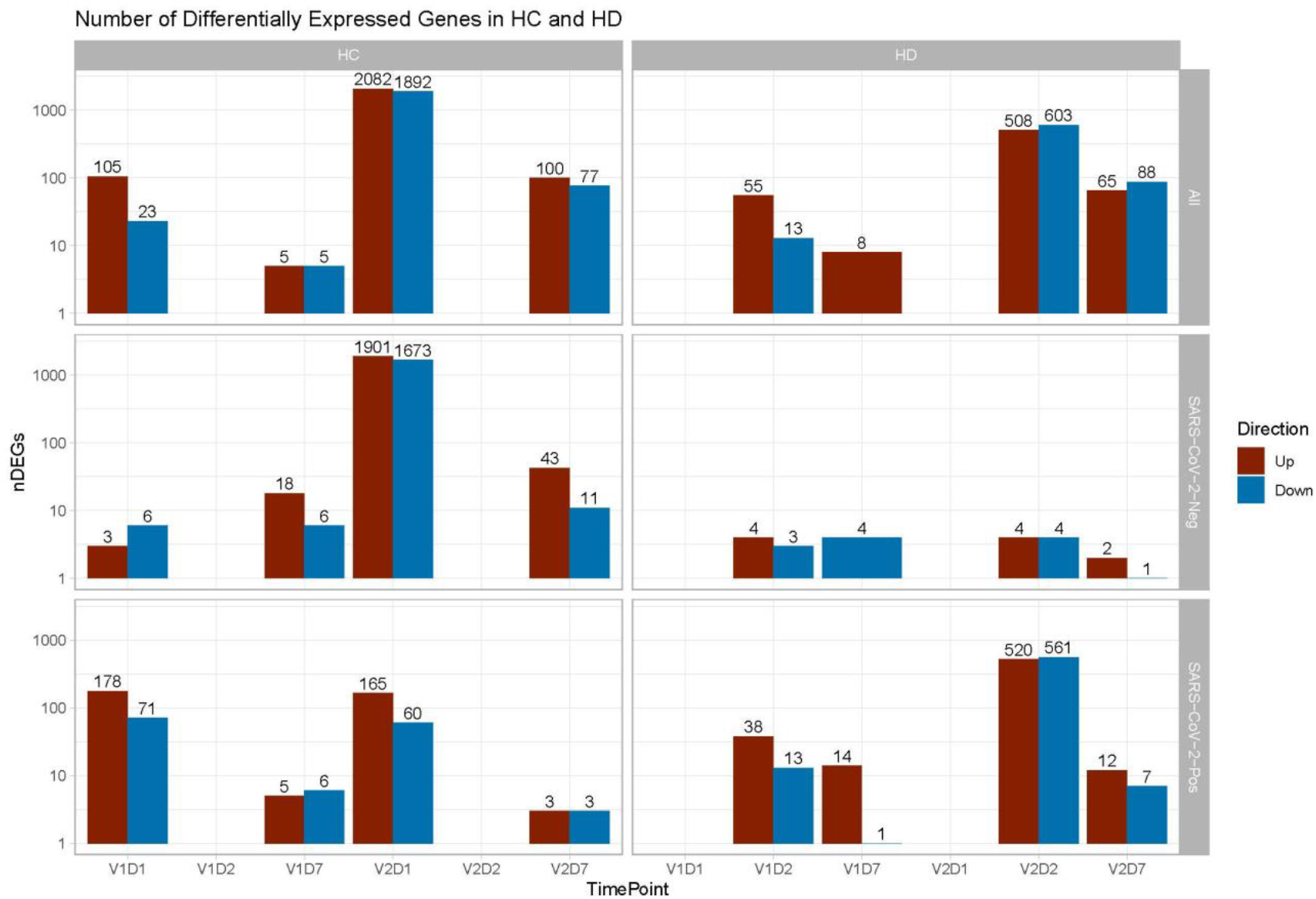
Differentially expressed genes (DEGs) increased after second vaccination dose compared to first, and at Day 1 and 2 (D1/D2) compared to Day 7 (D7) for both controls (HC) and maintenance hemodialysis (HD). Number of DEGs at each time point is displayed on a log scale, with DEGs for HC shown for D1 and D7 compared to pre-vaccination time point (D0), and DEGs for HD shown for D2 and D7 compared to D0. DEGs are shown independently of SARS-CoV-2 history **(Top)**, for analysis of only subjects with no prior SARS-CoV-2 history **(Middle)**, and for analysis of only subjects with prior SARS-CoV-2 history **(Bottom**). The DESeq2 R package was used to identify genes that were differentially expressed at each time point after vaccination for each subject group (p < 0.05, FDR-adjusted).

Direct comparison of gene expression between HC and HD with no prior reported SARS-CoV-2 infection at V1D7 yielded five DEGs in HD versus HC including increased expression of chemokine CCL19 in HD (p < 0.05, FDR-corrected). Comparison of these same groups at V2D7 yielded 18 DEGs including increased expression in HD of TIA1, which encodes a granule-associated protein expressed in cytolytic lymphocytes ^30^ and natural killer cells, and BH3, a pro-apoptotic Bcl-2 family member and mediator of lymphocyte apoptosis ^31^.

### Blood Transcription Module (BTM) Enrichment

BTM enrichment analysis of subjects without SARS-CoV-2 history reveals the vaccine-induced progression of various immune processes at each time point after vaccination (Figure 2). Following V1, HC demonstrate early (V1D1) enrichment of 29 BTMs, with substantial upregulation of monocyte and antiviral IFN activity (**TableS1**). The immune response transitions to V1D7 enrichment of four BTMs including significant T cell activation and downregulation of monocytes (**TableS2**). Following V2, HC demonstrate early (V2D1) enrichment of 82 BTMs, with substantial upregulation of innate antiviral activity, similarly to V1D1 (**TableS3**). The immune response transitions to V2D7 enrichment of ten BTMs, with significant upregulation of plasma cells and immunoglobulins (**TableS4**).

**Figure 2.**
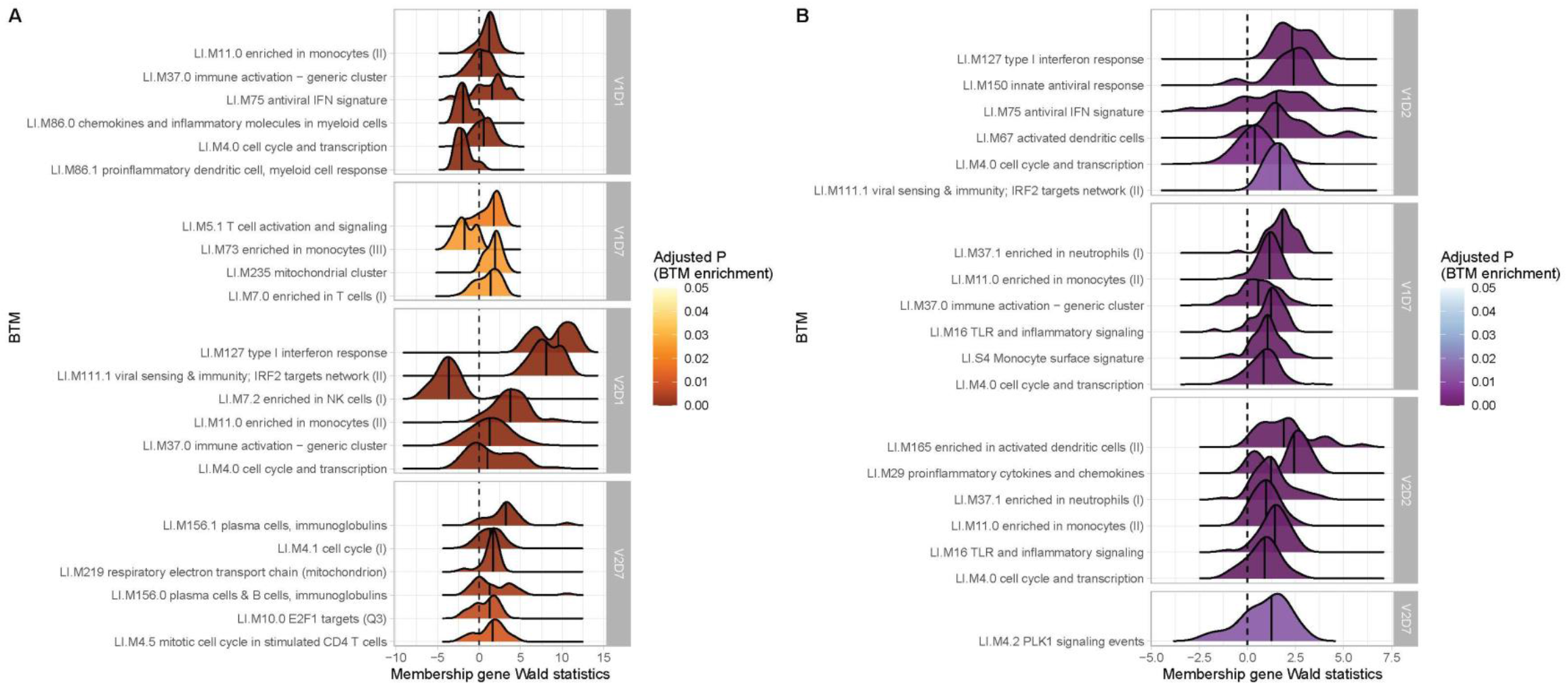
Controls (HC) and maintenance hemodialysis subjects (HD) with no SARS-CoV-2 history demonstrate differing longitudinal enrichments of blood transcription modules (BTMs). **(A)** The most significantly enriched BTMs are shown (up to six) for Day 1 (D1) and Day 7 (D7) after each vaccination dose (V1, V2) in HC with no prior infection with SARS-CoV-2 (p < 0.05, FDR-adjusted). Density plots for each BTM represent Wald statistics from DESeq2 analysis for each membership gene, thereby representing increased or decreased expression per gene at each time point compared to baseline (V1D0 or V2D0). **(B)** Similarly to (A), the most significantly enriched BTMs for Day 2 (D2) and Day 7 (D7) in HD are shown.

In contrast, HD demonstrate early (V1D2) enrichment of 12 BTMs after the first vaccination dose, most significantly involving upregulation of innate antiviral responses (**Figure 2, TableS5**). The immune response transitions to V1D7 enrichment of 17 BTMs, with substantial upregulation of myeloid modules (**Figure 2, TableS6**). The V1D7 positive enrichment of monocyte/myeloid modules in HD contrast the negative enrichment of these modules in HC (**Figure 2, Figure S1**). Following the second vaccination dose, HD demonstrate early (V2D2) enrichment of 27 BTMs most significantly involving upregulation of dendritic cell activity and proinflammatory cytokines and chemokines (**Figure3, TableS7**). The immune response progresses to V2D7 enrichment of one BTM: PLK signaling events (**Figure 2**).

**Figure 3.**
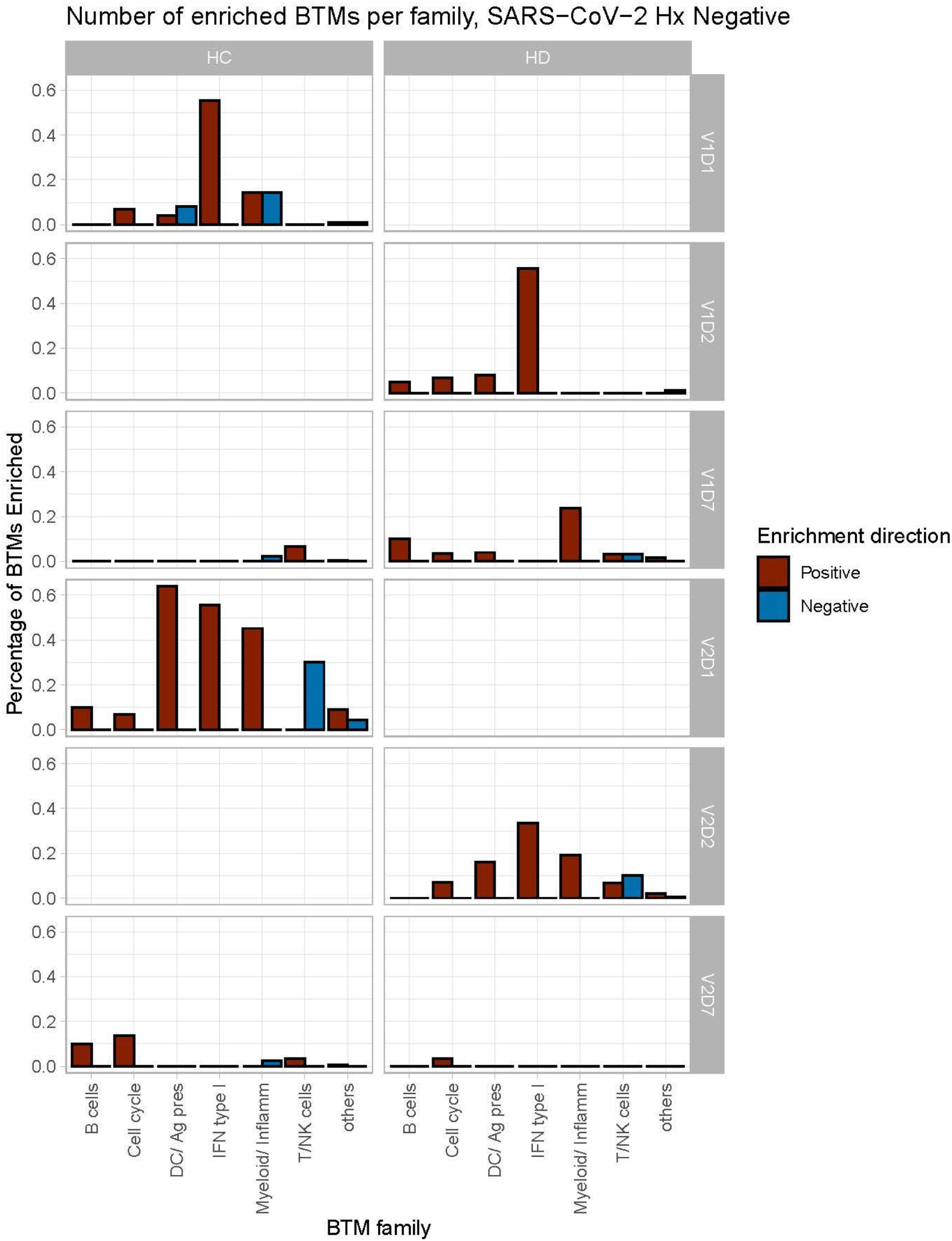
Controls (HC) and maintenance hemodialysis subjects (HD) demonstrate differing time courses of blood transcription module (BTM) enrichment after each vaccination dose. Percentage of BTMs in each BTM family with significant enrichment at each time point after each vaccination dose (V1, V2) for Day 1 (D1) and Day 7 (D7) in HC, and Day 2 (D2) and D7 for HD in subjects with no prior infection with SARS-CoV-2 (p < 0.05, FDR-adjusted). Direction of enrichment was determined using the median Wald statistic from DESeq2 analysis for each BTM membership gene, thereby representing overall increased or decreased expression of membership genes at each time point compared to baseline (V1D0 or V2D0).

**Figure S1.**
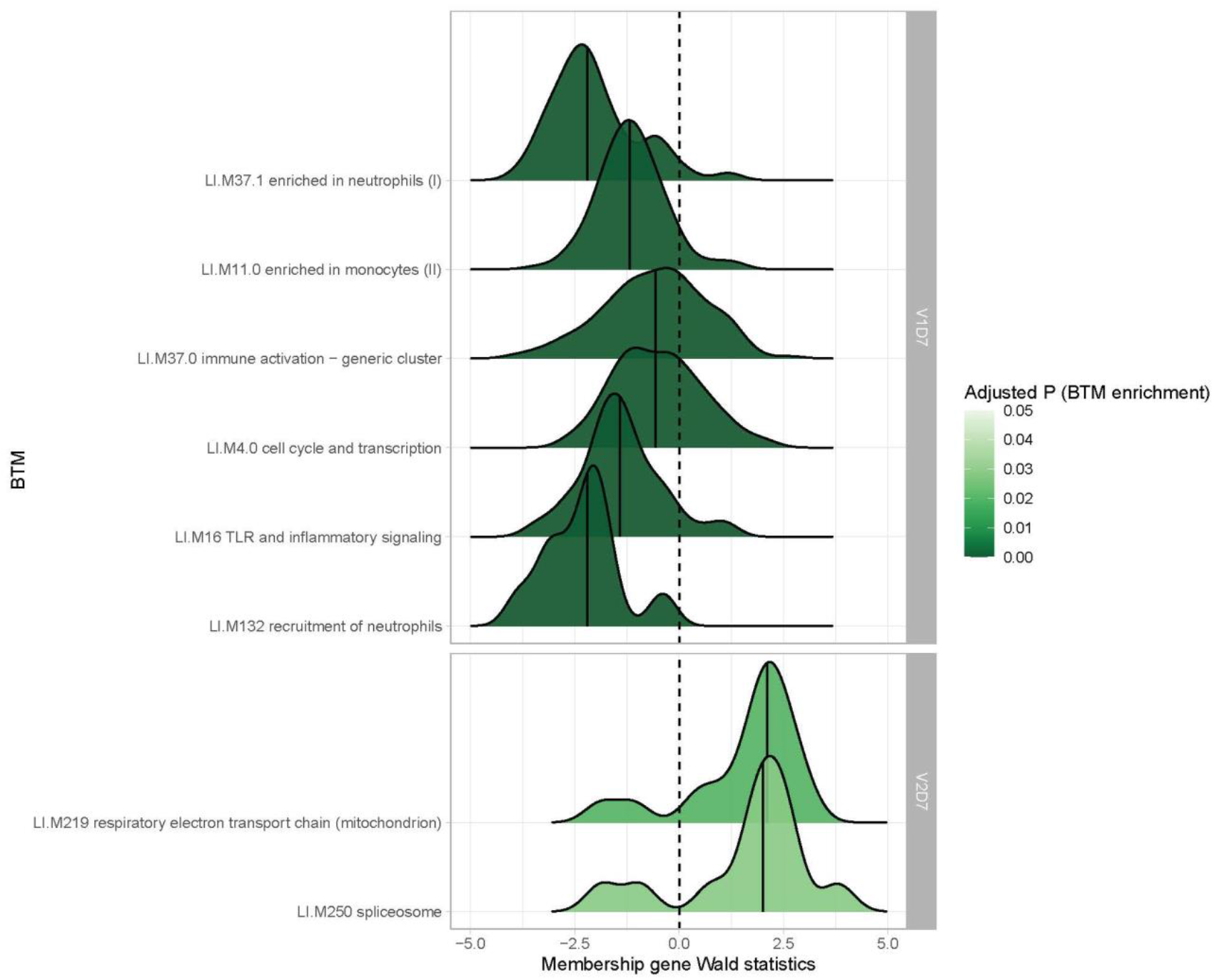
Hemodialysis patients (HD) without prior SARS-CoV-2 infection show increased myeloid activity at V1D7 and decreased metabolic activity at V2D7 compared to controls (HC). The most differentially enriched blood transcription modules (BTMs) between HC and HD with no prior infection with SARS-CoV-2 are shown (p < 0.05, FDR-adjusted) at V1D7 and at one week after second vaccination dose (V2D7). Density plots for each BTM represent Wald statistics from DESeq2 analysis for each membership gene per BTM, with positive Wald statistics indicating increased expression in HC compared to HD.

While there were no significant BTM enrichments in HC with positive SARS-CoV-2 history, most likely due to the insufficient number of subjects, BTM enrichments for HD with positive SARS-CoV-2 demonstrated notable upregulation of plasma cell activity at V1D7. This contrasts with V1D7 for HD with negative SARS-CoV-2, which show primary enrichment of myeloid BTMs (**Figure 2**). The remainder of these enrichments are shown in **Figure S2**.

**Figure S2.**
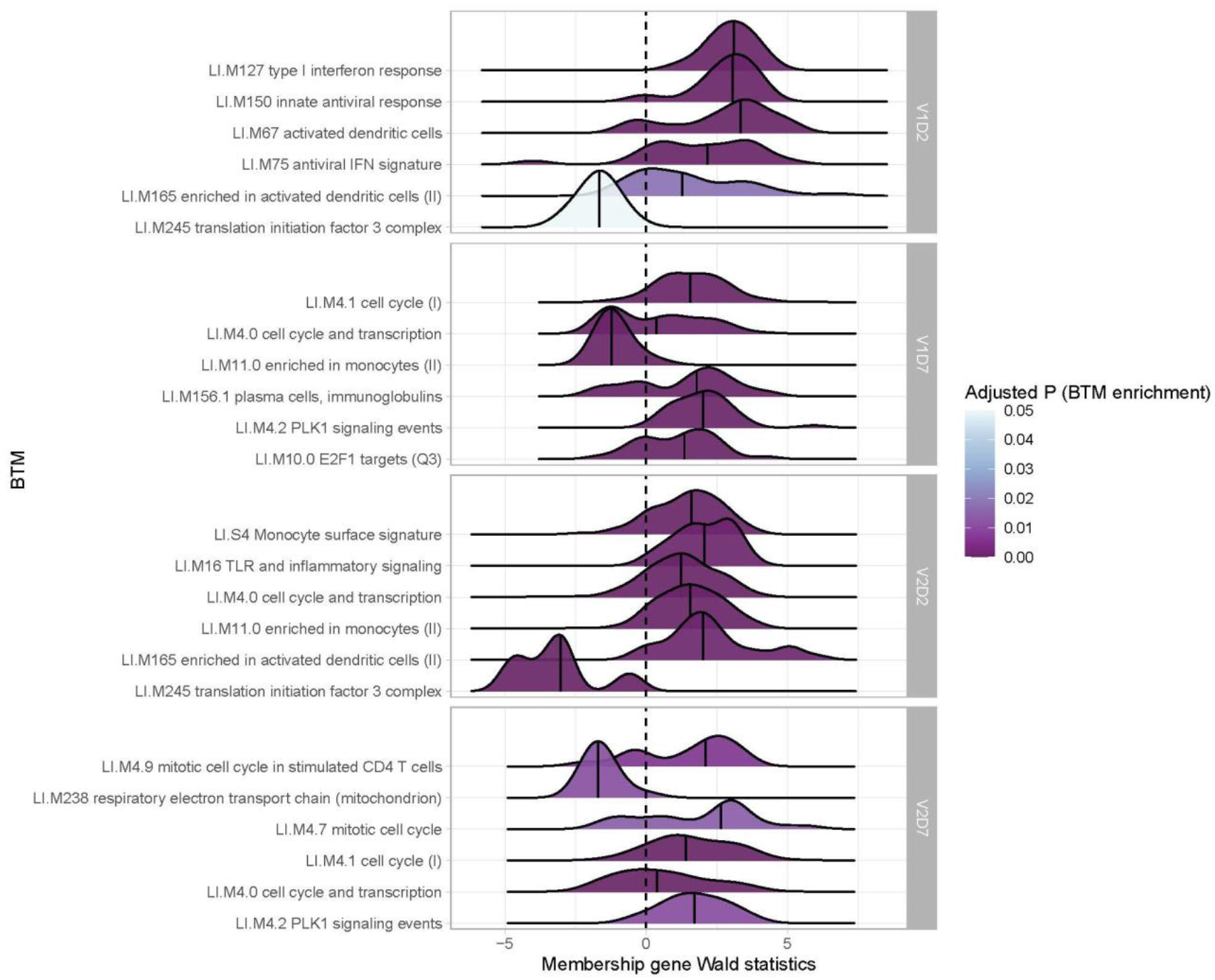
Hemodialysis patients (HD) with prior SARS-CoV-2 infection show increased expression of innate and adaptive immune blood transcription modules (BTMs) post-vaccination. The most significantly enriched BTMs are shown (up to six) for Day 2 (D2) and Day 7 (D7) after each vaccination dose (V1, V2) in HD with prior infection with SARS-CoV-2 (p < 0.05, FDR-adjusted). Density plots for each BTM represent Wald statistics from DESeq2 analysis for each membership gene, thereby representing increased or decreased expression per gene at each time point compared to baseline (V1D0 or V2D0).

Summary enrichments using BTM families show many positive early V1 enrichments of Type 1 IFN activity that dissipate by V1D7 in both HC and HD (**Figure 3**). However, HC show early positive and negative enrichments of myeloid/inflammatory family activity that dissipate by V1D7, while HD show many early positive enrichments of myeloid/inflammatory family activity that persist and increase at V1D7. Following V2, HC show early predominance of dendritic cell (DC)/antigen presenting cell (APC), IFN Type I, and myeloid/inflammatory family activity transitioning to B cell and cell cycle activity at V2D7, while HD show predominant early IFN type I family activity transitioning to just one detectable cell cycle module enrichment.

### Antibody Binding and Neutralization Assay Response

We next aimed to assess immune protection conferred by the vaccine through quantification of anti-spike IgG antibodies and functional assessment of neutralizing antibodies. All subjects demonstrated an increase in anti-spike IgG at V2D7, with titers for all subjects except one still elevated above baseline at six months. The exception was one HD subject with prior SARS-CoV-2 infection who demonstrated the highest baseline titers of all subjects prior to vaccination. Both HC and HD subjects demonstrated a statistically significant increase in anti-spike IgG and neutralization activity (ID50) from V1D0 to V2D7 (p < 0.001), followed by an expected decrease at M6 from V2D7 levels (p < 0.001) (**Figure 4**). Despite this decrease, M6 titers were still increased compared to baseline (p< 0.001).

**Figure 4.**
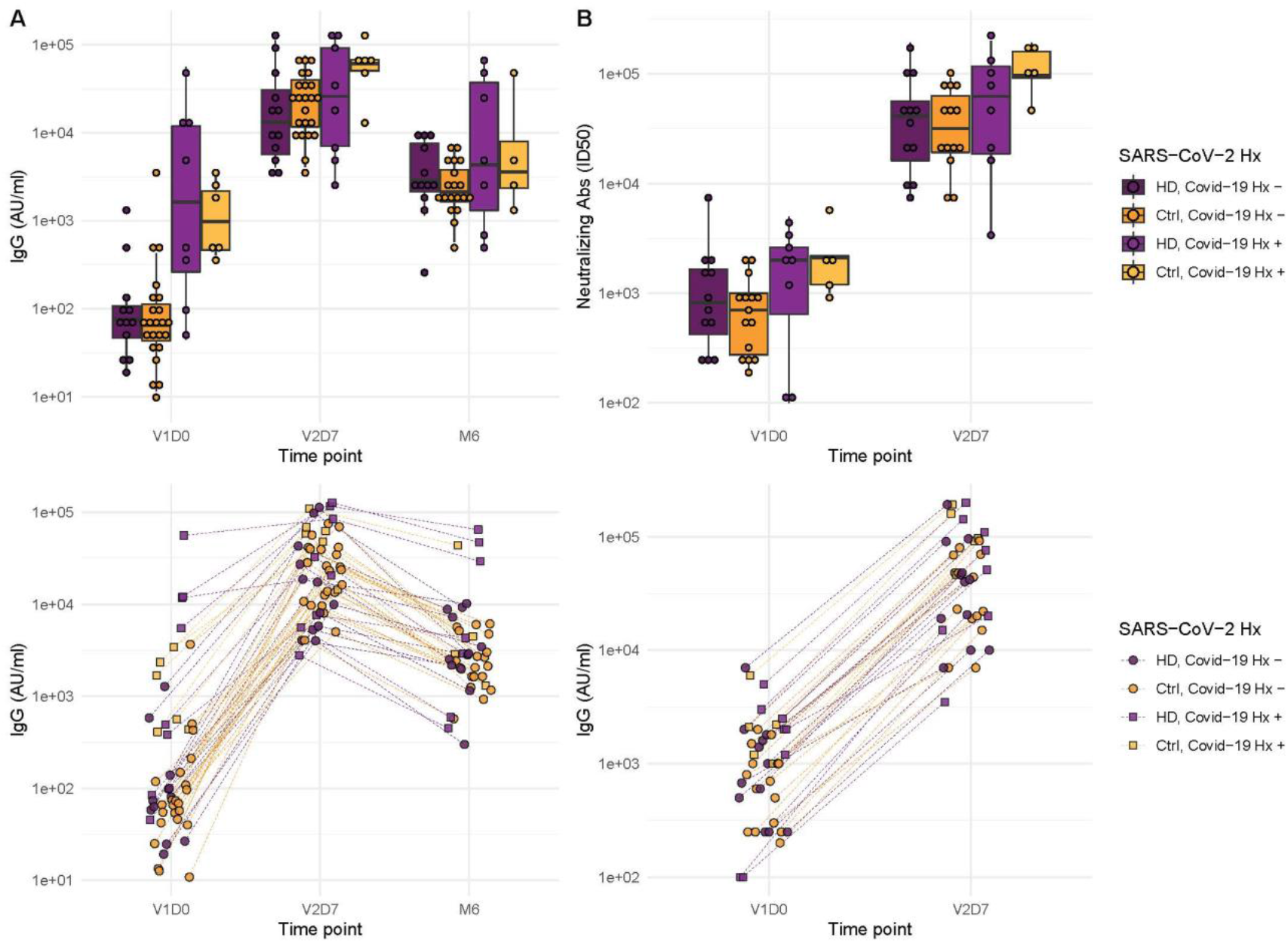
Antibodies significantly increased in controls and maintenance hemodialysis (HD) one week after the second vaccination dose (p < 0.001) and six months after initial vaccination (p < 0.001) with the BNT162b2 mRNA COVID-19 vaccine. (**A**) Anti-spike IgG levels in controls and HD subjects with and without prior SARS-CoV-2 history before vaccination (V1D0), one week after second vaccination dose (V2D7), and six months after initial vaccination (M6). (**B**) Antibody neutralization activity (ID50) in controls and HD subjects with and without prior SARS-CoV-2 history at V1D0 and V2D7.

Higher anti-spike IgG at V2D7 was significantly predicted by higher pre-vaccination anti-spike IgG, control group assignment, and younger age (p <0.01, p<0.05, p<0.05, respectively), while gender, race, and ethnicity were not. Higher anti-spike IgG at M6 was significantly predicted by higher V2D7 anti-spike IgG (p < 0.001), with no additional predictive value conferred by SARS-CoV-2 history, subject group, age, gender, race, or ethnicity. Higher neutralization activity (ID50) at V2D7 was significantly predicted by higher pre-vaccination ID50, with no additional predictive value conferred by subject group, age, gender, race, and ethnicity.

### Transcriptomic and clinical predictors of antibody binding response in HD

Linear models to predict anti-Spike IgG at V2D7 and at M6 in HD using enriched BTMs, controlling for SARS-CoV-2 history, identified BTM predictors at all time points except for V1D2. Of the 18 enriched BTMs at V1D7, increased expression (from V1D0) of “LI.M156.1 plasma cells, immunoglobulins” was predictive of higher anti-spike IgG at V2D7 (p < 0.05, FDR-corrected), controlling for SARS-CoV-2 history. Of the 30 enriched BTMs at V2D2, increased expression of 18 BTMs was predictive of higher anti-Spike IgG at V2D7 (p < 0.05, FDR-corrected). These include innate immune, antigen presentation, and T cell pathways (**Figure 5**). Increased expression of “LI.M4.2 PLK1 signaling events” at V2D7 compared to V2D0, which was the only enriched module at this time point for HD subjects with no SARS-CoV-2 history, was predictive of higher anti-spike IgG at both V2D7 and M6 (p < 0.05).

**Figure 5.**
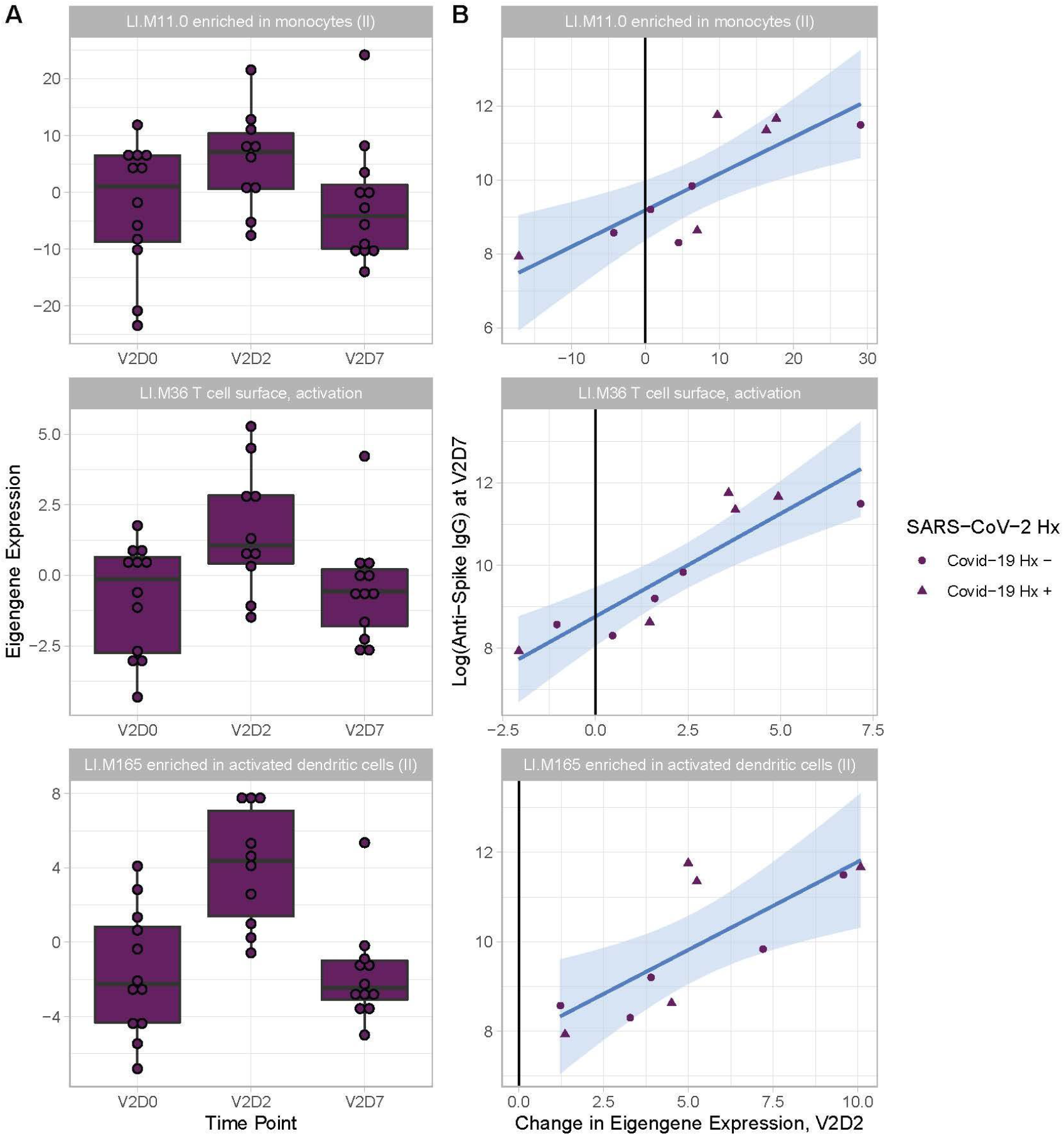
Increased expression of multiple Blood Transcription Modules (BTMs) at V2D2 is predictive of higher anti-spike IgG at V2D7. Of 30 enriched BTMs at V2D2, increased expression of 18 BTMs is predictive of increased anti-spike IgG at V2D7 (p< 0.05, FDR-corrected), controlling for SARS-CoV-2 history. Predictive pathways include innate immune, antigen presentation, and T cell pathways. Three examples are shown.

Linear models to predict anti-Spike IgG at V2D7 and at M6 In HD using clinical predictors yielded significant baseline and post-vaccination predictors. Baseline ferritin levels in the intermediate range (200 – 2000 ng/ml) were associated with higher anti-spike IgG at V2D7 and M6 (p < 0.01, p < 0.05), controlling for history of SARS-CoV-2. URR, WBC counts, transferrin saturation, and hemoglobin were not significant predictors of antibody development. **Figure 6A** shows anti-spike IgG at V2D7 as a function of baseline ferritin levels, identifying the intermediate range of ferritin which has previously been associated with lowest all-cause mortality ^20^

**Figure 6.**
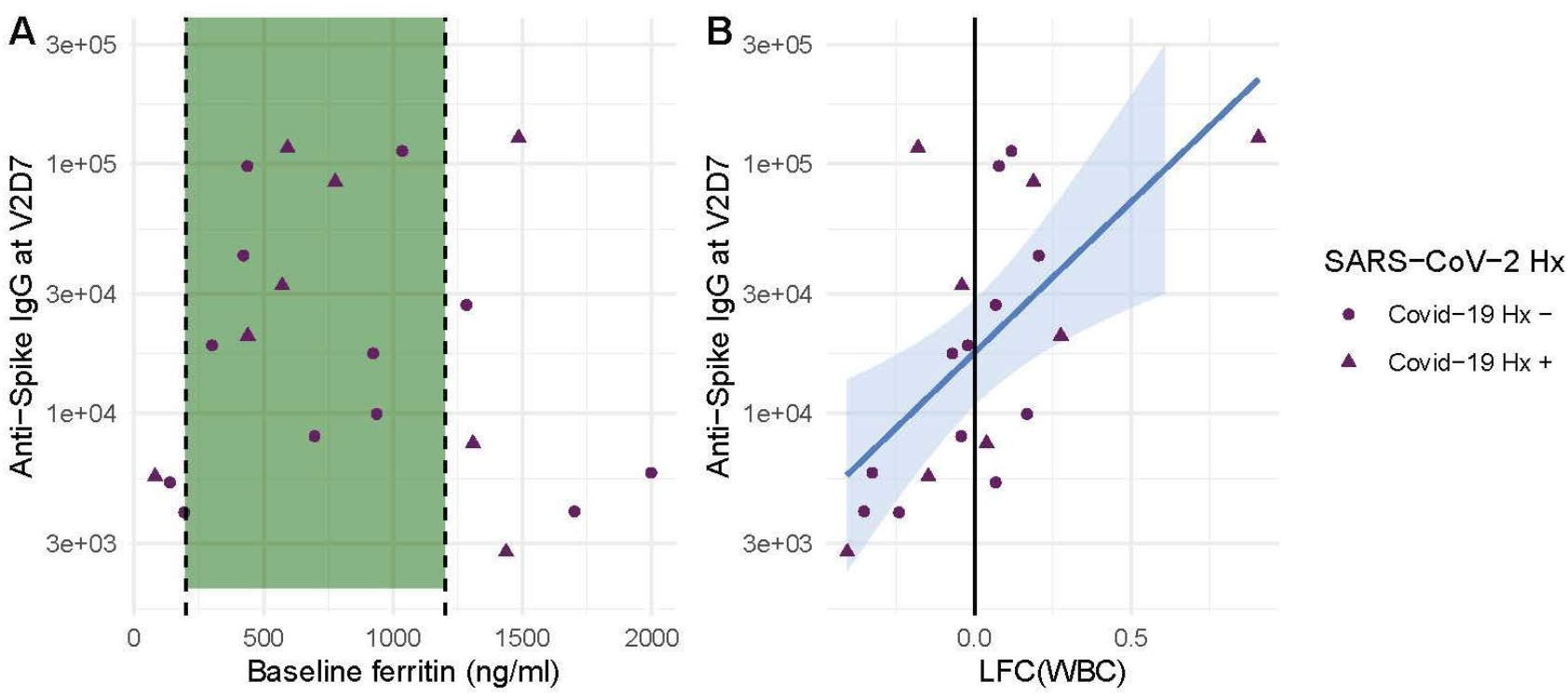
Baseline ferritin level and post-V1 white blood cell count (WBC) are clinical predictors of post-V2 antibody responses in maintenance hemodialysis patients (HD). (A) Ferritin levels associated with lowest all-cause mortality predict the development of higher anti-spike IgG after vaccination at V2D7 (p < 0.01) and M6 (not shown, p < 0.05) in maintenance HD patients. Dashed vertical lines indicate the intermediate range of ferritin (200-1200 ng/ml) associated with lowest all-cause mortality ^20^. (B) Increased WBC after first vaccination dose is predictive of anti-spike IgG titers after vaccination at V2D7 (p < 0.01) and M6 (not shown, p < 0.05) in maintenance HD patients. Points with negative log-fold change of white blood cell counts (LFC(WBC)) and positive LFC(WBC) represent a decrease and increase, respectively, in WBC from baseline labs.

The LFC of WBCs from baseline after the first vaccination dose was significantly predictive of antibody titer levels at both V2D7 (p <0.01) and M6 (p < 0.05), controlling for SARS-CoV-2 history and number of days after vaccination that labs were collected (**Figure 6B**). The predictive value of LFC of WBCs is predominantly driven by increased lymphocyte counts; LFC of absolute lymphocyte counts was predictive of V2D7 (p < 0.01) and M6 (trend-level, p = 0.056) antibody titers, controlling for initial antibody titers and date of clinical labs.

## Discussion

Our results demonstrate differing expression of BTMs and differing time courses of immune responses to the BTN162b2 mRNA COVID-19 vaccination in maintenance hemodialysis subjects (HD) compared to controls. Controls demonstrated expected transitions from early Type I interferon and myeloid activity to T cell activity after the first vaccination dose (**Figure 2, Figure 3**). The predominant positive enrichment of T cell modules in controls at one week after the first vaccination dose (V1D7) was contrasted with predominant positive enrichment of myeloid modules in HD at V1D7. Interestingly, HD showed prolonged upregulation of myeloid activity at V1D7, while controls showed downregulation of these modules at V1D7 (**Figure 2, Figure S1**). Overall, these observations indicate delayed resolution of innate myeloid responses in the HD cohort.

Following the second vaccination dose, both groups demonstrated early enrichment of innate immune modules, with HC alone transitioning to a plasma cell response by V2D7 (**Figure 3**). Direct group comparisons at V2D7 did not show differences of plasma cell response, but metabolic activity was decreased in HD compared to controls (**Figure S1**). Further, HD demonstrated increased V2D7 expression compared to controls of pro-apoptotic Bcl-2 family member BH3, a mediator of lymphocyte apoptosis. A prior study showed accelerated *in vitro* apoptosis of lymphocytes in uremia, with a particularly pronounced effect on B cells, mediated by dysregulation of Bcl-2. These results suggest a state of heightened cellular stress in HD after vaccination leading to increased apoptotic signaling.

Despite differing transcriptomic time courses in the two group, our results demonstrate significant elevation of anti-spike IgG titers after two doses of BNT162b2 mRNA COVID-19 vaccination in both HD and controls. HD demonstrated only a slight decrease of IgG levels at V2D7 when controlling for SARS-CoV-2 history (p < 0.05) and no statistically significant difference at six months. Prior studies comparing short-term antibody response to BNT162b2 mRNA COVID-19 vaccination in HD versus controls find antibody response rates of 84-96% in HD after two vaccination doses, but with lower mean IgG levels compared to controls ^16–19,32–34^. Notably, the HD population studied here is younger and more racially and ethnically diverse. The average age of HD cohorts in prior studies was predominantly in the 60s, compared to an average age of 54 in our study. Jahn et al. found in a subset analysis that HD patients under 60 years of age responded equally to healthy controls, suggesting an interaction between increasing age and less effective antibody response in HD patients ^17^.

HD subjects with documented SARS-CoV-2 infection prior to vaccination had wider variance of antibody titers at all time points in this study, with two subjects demonstrating V1D0 antibody titer levels similar to that of uninfected subjects. These two subjects consistently had the lowest titer levels at V2D7 and M6 within the group of previously infected subjects and amongst the lowest titers across all subjects. One subject is the oldest enrolled patient, and both are diagnosed with hyperlipidemia.

Given previously and presently demonstrated the wider variance of protective immune responses in HD and altered interactions with risk factors including age, it is valuable to identify predictors of the strength of immune response to vaccination in this population. We identified both transcriptomic and clinical predictors of anti-spike IgG development at both V2D7 and six months after the second vaccination dose (M6). Increased gene expression of blood transcription modules (BTMs) including monocyte activity, dendritic cell and antigen presentation activity, IFN type I activity, and T cell activation two days after the second vaccination dose (V2D2) in HD were predictive of V2D7 anti-spike IgG. Additionally, increased expression of PLK1 signaling events, indicating increased cell cycle activity, at V2D7 was predictive of V2D7 and M6 anti-spike IgG. Clinically, serum ferritin values in the intermediate range at baseline predicted stronger anti-spike IgG development. A prior study of 58,058 maintenance HD subjects found serum ferritin levels between 200 and 1200 ng/ml to be associated with lower all-cause mortality, due to ferritin <200 ng/ml representing low iron status, and >1200 ng/ml representing a hyper-inflammatory state due to ferritin’s status as an acute phase reactant ^20^. Iron deficiency has been linked to impaired immune response and vaccine efficacy in other infections, while inflammation induces macrophage release of the heavy chain component of ferritin, FTH, which has been reported to inhibit lymphocyte proliferation and function ^34,35^. Additionally, increased LFC in WBC count 1-3 weeks after vaccination was predictive of higher antibody titers.

Our study is limited by different early time points between controls and HD (Day 1 vs Day 2 after each vaccination dose) and by sample size, particularly when subdividing SARS-CoV-2 history. The smaller sample size additionally limits our ability to characterize differential immune pathways in clinical subsets of the dialysis population, such as those with low, medium, and high baseline ferritin levels. Future studies are needed for more comprehensive characterization of the immune pathway recruitment in response to the Covid-19 vaccinations in this population. The Covid-19 mRNA vaccines are proving more efficacious than other vaccines in the ESRD population; for example, while more than 90% of patients without chronic kidney disease develop protective antibodies against hepatitis B after vaccination, only 50-60% of patients with ESRD seroconvert ^9^. One explanation is that, in mRNA vaccines, the mRNA both encodes the viral antigen and acts as an adjuvant due to its innate immunostimulatory properties; the mRNA is recognized by endosomal and cytosolic innate sensors upon cell entry, resulting in cellular activation and production of type I interferons and other inflammatory mediators ^36^. This elevated innate immune stimulus could overcome immune desensitization in ESRD, evidenced by diminished TLR4 expression on monocytes ^37^ and downregulation of activating receptors on natural killer cells in this population ^38^. If so, the mRNA vaccine delivery vehicle could prove particularly valuable in vaccine development for ESRD and HD going forward.

Overall, we demonstrate differing time courses of immune responses to the BTN162b2 mRNA COVID-19 vaccination in maintenance hemodialysis subjects (HD) and identify transcriptomic and clinical predictors of anti-Spike IgG titers in HD. Given the efficacy of this vaccination in our HD cohort, the differential transcriptomic responses likely represent immune alterations with sub-clinical effects on humoral immune response to the BTN162b2 mRNA COVID-19 vaccine. Our results warrant further characterization of the immune dysregulation of ESRD and of immune biomarkers that underlie efficacious immune responses to vaccination in this population.

## Data Availability

Sequencing data have been deposited in GEO under accession code GSE209985.

